# Zero-Cost Digital Automation for Scalable Assessment in a National Mechanical Ventilation Curriculum

**DOI:** 10.64898/2026.09.11.26362682

**Authors:** Amarpreet K. Ahluwalia, Khoi N. Tran, Bashar S. Staitieh, Michael B. Keller, Junfeng Sun, Burton W. Lee, Nitin Seam, the Critical Care Education Research Consortium

## Abstract

**Background:** As medical education transitions toward competency-based models, longitudinal programs emphasizing deliberate practice and repeated assessment are essential for mastery of complex concepts. Frequent formative assessment requirements, limited expert pools, and growing cohorts that outpace traditional infrastructure create substantial scalability constraints. Preceptorial programs, in which learners demonstrate mastery to expert educators, are particularly vulnerable. As enrollment expands, administrative demands for portfolio tracking, examination logistics, and case development fall disproportionately on the volunteer expert educators who sustain them.

**Objective:** We sought to implement a unified, zero-cost digital infrastructure to support scalable formative and summative assessment of a multi-center year-long mechanical ventilation preceptorial program while minimizing administrative burden for expert educators.

**Methods:** We implemented two complementary digital systems built on the Google ecosystem, requiring no institutional licensing or IT support. For formative tracking, we first deployed a standardized WP with defined milestone deadlines and a zero-cost automated tracking and reminder system. Completion of the fall milestone was compared between Fall 2024 (manual educator-dependent tracking and reminders) and Fall 2025 (standardized requirements plus automated tracking). For summative assessment, we also deployed an automatically graded multiple-choice case-based waveform examination (CBWE), operationalizing an expert-defined partial-credit scoring rubric.

**Results:** There was a 7.7-fold increase in the total number of waveforms submitted (194 images in Fall 2024 to 1498 images in Fall 2025). Portfolio submission rate rose from 45.6% (31/68 learners) in Fall 2024 to 68.5% (63/92 learners) in Fall 2025. Among learners who submitted their portfolios, the proportion submitting all required waveforms increased from 29.0% to 55.6%. Proof-of-concept automated CBWE grading confirmed scoring accuracy, eliminated 8.5 hours of manual grading per grader per cycle, generated item-level analytics, and demonstrated capacity for immediate individualized learner feedback.

**Conclusion:** A unified, zero-cost digital infrastructure addressed parallel administrative and content scarcity constraints limiting scalability of a national MV preceptorial program. By reallocating administrative burden away from expert faculty, these tools preserve the high-fidelity teaching relationship at the core of the preceptorial model while enabling it to operate at scale, ultimately providing a replicable model for competency-based assessment in diverse training environments.

## Introduction

As medical education transitions to more competency-based models, there will be increased emphasis on deliberate practice with repeated assessments to develop mastery of specialty and subspecialty-specific foundational competencies.[1-3] However, frequent formative assessment requirements, limited expert pools, and growing cohorts pose substantial barriers to scalability of competency-based medical education, threatening sustainability of the very programs designed to meet these educational demands. Low-cost digital tools represent a potential replicable model for competency-based programs operating across diverse environments with limited centralized resources, given limited funding for GME. While multi-center collaboration can help scale competency-based education beyond these local constraints, heterogeneous digital tools and security policies, along with the need to preserve assessment confidentiality, pose significant barriers to deployment.

Management of mechanical ventilation (MV) is a foundational competency for Pulmonary and Critical Care Medicine (PCCM) and Critical Care Medicine (CCM) practitioners and a core competency for PCCM and CCM fellows.[4-6] Mastery requires real-time integration of respiratory physiology, waveform interpretation, and recognition and management of patient-ventilator asynchronies.[7, 8] Despite its central importance, there is no standardized curriculum or assessment framework for MV training in the United States, resulting in substantial variability in educational approaches across fellowship programs.[4, 9, 10]. Deficits in ventilator waveform interpretation may have meaningful clinical consequences as higher rates of unrecognized asynchrony are associated with increased tracheostomy and mortality.[11, 12] Prior studies demonstrate that practicing intensivists correctly identify only approximately one-third of common patient-ventilator asynchronies.[13]

A structured MV course significantly improved fellows’ identification and management of ventilator asynchronies compared with traditionally trained peers.[7] However, subsequent longitudinal evaluation revealed that such knowledge gains decayed over time unless reinforced by deliberate practice.[14] The MV preceptorial is a one-on-one, teach-the-expert model based on the Oxford tutorial tradition, in which learners synthesize and demonstrate mastery of core concepts to an expert educator rather than passively receiving instruction from one.[15] As part of this program, learners collect deidentified ventilator waveform images from clinical practice, annotate them with physiologic interpretation, and provide clinical reflection in a waveform portfolio (WP), a process that engages higher-order cognitive skills aligned with Bloom’s taxonomy and produces a durable teaching artifact that can be revisited throughout training.

As enrollment in the preceptorial program expanded to PCCM and CCM fellows and faculty across the country, the model, intentionally designed to leverage dedicated expert time for high-fidelity teaching, increasingly strained the administrative infrastructure required to support it. In programs of this kind, administrative tasks spanning registration of learners, longitudinal tracking of performance across modules, and assessment logistics can consume hundreds of hours of total faculty time per program cycle. Manual portfolio tracking and individualized examination grading placed disproportionate logistical demands on the limited pool of expert faculty volunteers. A written case-based waveform examination (CBWE), previously administered on paper across sites, required physical distribution and centralized return for grading, precluding timely learner feedback, a foundational element of competency-based assessment. Simultaneously, the scarcity of vetted, deidentified MV cases limited the program’s ability to expand or refresh its assessment content of clinical waveforms over time.

We sought to preserve the integrity of the preceptorial model while offloading its administrative burden through a unified, zero-cost digital infrastructure accessible across all participating institutions, one designed to improve formative assessment completion, enable automated summative grading with individualized feedback, and leverage learner-generated waveform portfolios as a pipeline for future examination content. Critically, we intentionally avoided proprietary learning management systems, which vary in availability across institutions and may create participation barriers for programs with limited resources. By utilizing the Google ecosystem and allowing learners to participate via personal Gmail accounts, the infrastructure required no institutional IT support, licensing, or procurement, lowering the barrier to participation for fellows and practicing intensivists nationwide.

## Methods

We developed two complementary digital systems: 1) a WP submission and tracking system and 2) an automated CBWE grading system to support scalable formative and summative assessment within a national training program. Using a pre-post design, timely completion of fall portfolio requirements was compared between Fall 2024 (manual tracking and reminders) prior to implementation of the digital systems and Fall 2025 (standardized requirements plus automated tracking and reminders). Participants included CCM and PCCM fellows and practicing attending physicians trained in CCM or PCCM. This study was deemed exempt from review under a multisite IRB protocol.

The intervention comprised two components: 1) an expert-vetted, standardized waveform portfolio structure with defined fall and spring milestone deadlines, and 2) a zero-cost automated tracking and reminder system. Learners accessed the system via personal Gmail accounts, ensuring accessibility without reliance on proprietary learning management systems, institutional IT infrastructure, or licensing.

### Standardization of Waveform Submissions and Milestones

A standardized submission deck grouped deidentified waveform photographs into three major categories: normal waveforms, maneuvers, and asynchronies, with example submissions and recommended annotations provided to clarify expectations. Three authors (A.A., B.S., N.S.) reviewed and consolidated historical waveform requirements. Additional advanced maneuvers and cases deemed of high educational value were incorporated into the updated submission list following review (Figure 1). Learners were permitted to submit the same waveform as evidence of multiple requirements if annotated appropriately. Submission requirements were organized into two milestone deadlines aligned with modules completed during the fall and spring semesters.

**Figure 1.**
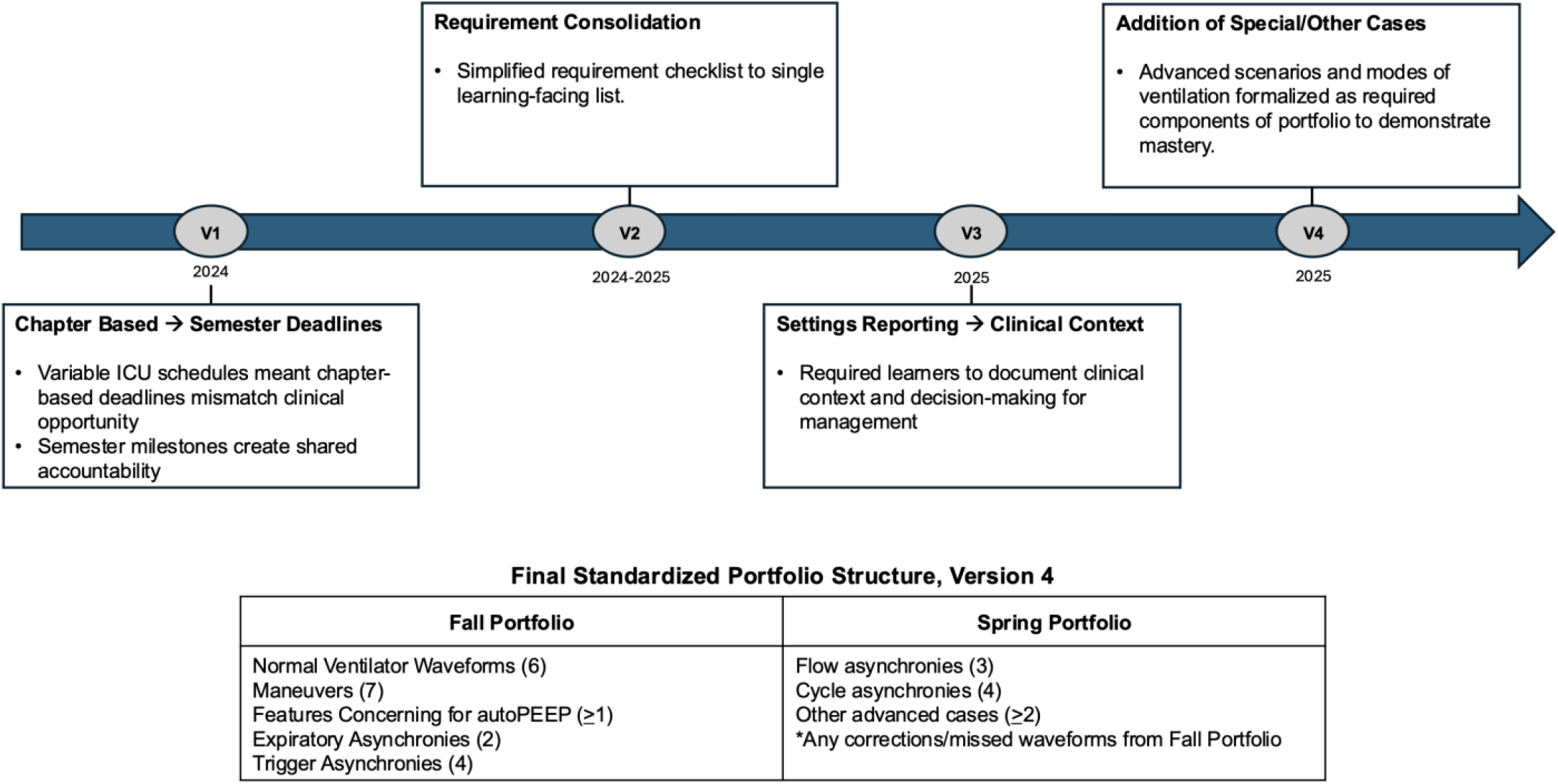
Iterative development of the standardized waveform portfolio (WP) submission requirements. Top Panel: Timeline of major structural revisions to WP requirements across four iterative versions (V1, 2024; V2, 2024-2025; V3-V4, 2025), including a shift from chapter/module-based to semester-aligned milestone deadlines, consolidation of fluency tiers into a single required-item list, addition of a clinical-context/settings-reporting requirement, and formalization of previously optional advanced cases into a required “Other Cases” category. Bottom Panel: Final standardized Fall and Spring portfolio requirements (Version 4, 2025-2026), organized by waveform category.

Learners submitted deidentified waveform photographs compiled into the standardized template (Supplemental Figure 1). Prior to storage and any potential reuse, image metadata was stripped by program leadership, and submissions underwent expert review and cropping to ensure removal of any potentially identifying information. Access to submission folders was restricted to individual learners and course administrators. No protected health information was collected or stored within the system. Percentage completed scoring was capped at 100% regardless of total number of image submissions.

### Automated Tracker and Learner Nudge System

A zero-cost automated tracking infrastructure was developed using Google Sheets and Google Apps Script (Google, Mountain View, CA). Generative AI (ChatGPT 4o/5, Gemini 2.5/3) was used iteratively for code drafting and debugging (Supplemental Material).

The system maintained a centralized learner tracking sheet, automatically updated submission status, and generated individualized reminder emails to learners at predefined intervals. During the submission period (November-December 2025), weekly reminder emails were sent only to learners who had not yet submitted a portfolio. Weekly summary emails were sent to course leadership reporting aggregate portfolio submission status.

### Summative Case-Based Written Examination Grading Automation

The existing CBWE consists of five images depicting asynchronies and multiple-choice questions requiring multiple selections corresponding to identification and management of visualized asynchronies. An existing consensus scoring rubric includes full-credit, partial-credit, and penalty options for responses representing potentially harmful management decisions (Supplemental Figure 2). A zero-cost automated grading system was designed using Google Forms and Google Apps Script to operationalize this rubric while preserving the expert-defined assessment structure (Supplementary Information).

### Manual Grading Workflow and Workload Assessment

To estimate faculty grading workload, expert graders (A.A., N.S.) timed grading of six previously administered, deidentified waveform examinations. Examinations were selected to represent high-, middle-, and low-scoring learner performance (B.L.). Mean grading time per examination was used to estimate projected faculty grading workload for varying cohort sizes. Two authors (A.A., K.T.) independently verified automated scoring outputs against manually graded examinations to ensure fidelity to adjudicated scoring rubric.

### Study Outcomes

The primary outcome was timely submission of WP, defined as the proportion of learners submitting fall portfolio requirements by the fall milestone deadline, compared between the control cohort (manual tracking and reminder emails) and the intervention cohort (standardized portfolio requirements with automated tracking).

Secondary outcomes included: (1) estimated faculty grading workload under the existing manual examination process, (2) proof-of-concept demonstration of automated grading system functionality and scoring accuracy prior to prospective deployment, and (3) distribution of portfolio completion among submitters (proportion completing 100% of required waveforms; median percentage of required waveforms completed), assessed as an exploratory outcome.

### Data Analysis

Portfolio completion rates were compared between the control (Fall 2024) and intervention (Fall 2025) cohorts using R version 4.6.1 and Python. Categorical outcomes were compared between cohorts using Fisher’s exact test (two-sided). The distribution of percentage of required waveforms completed among submitters was compared using the Mann-Whitney U test. Given the pre-post observational design and potential for confounding between cohorts (including growth in enrollment and program maturity), between-cohort comparisons are presented as descriptive associations. Manual grading time was summarized using mean values across graders and examinations and used to estimate projected faculty grading workload for varying cohort sizes. Scoring accuracy was assessed by comparing assigned scores with the adjudicated scoring key.

## Results

### Waveform Portfolio Submission

In 2024-2025, a total of 68 learners (42 fellows, 26 attendings) from 19 unique institutions were enrolled; in 2025-2026, a total of 92 learners (59 fellows, 33 attendings) from 24 unique institutions were enrolled. Fall portfolio submission attempts were significantly higher in the automated-tracking cohort compared with the manual-tracking cohort (68.5% [63/92] vs 45.6% [31/68]; p=0.006). The proportion submitting all required waveforms increased from 29.0% to 55.6%, p = 0.017. The median completion of required waveforms rose from 94.7% (IQR 57.9-100.0%) to 100.0% (IQR 91.7-100%), Mann-Whitney p=0.003 (Table 1). A total of 26 learners in 2025-2026 cohort submitted additional non-required waveforms, which were only counted in total images received.

**Table 1.**
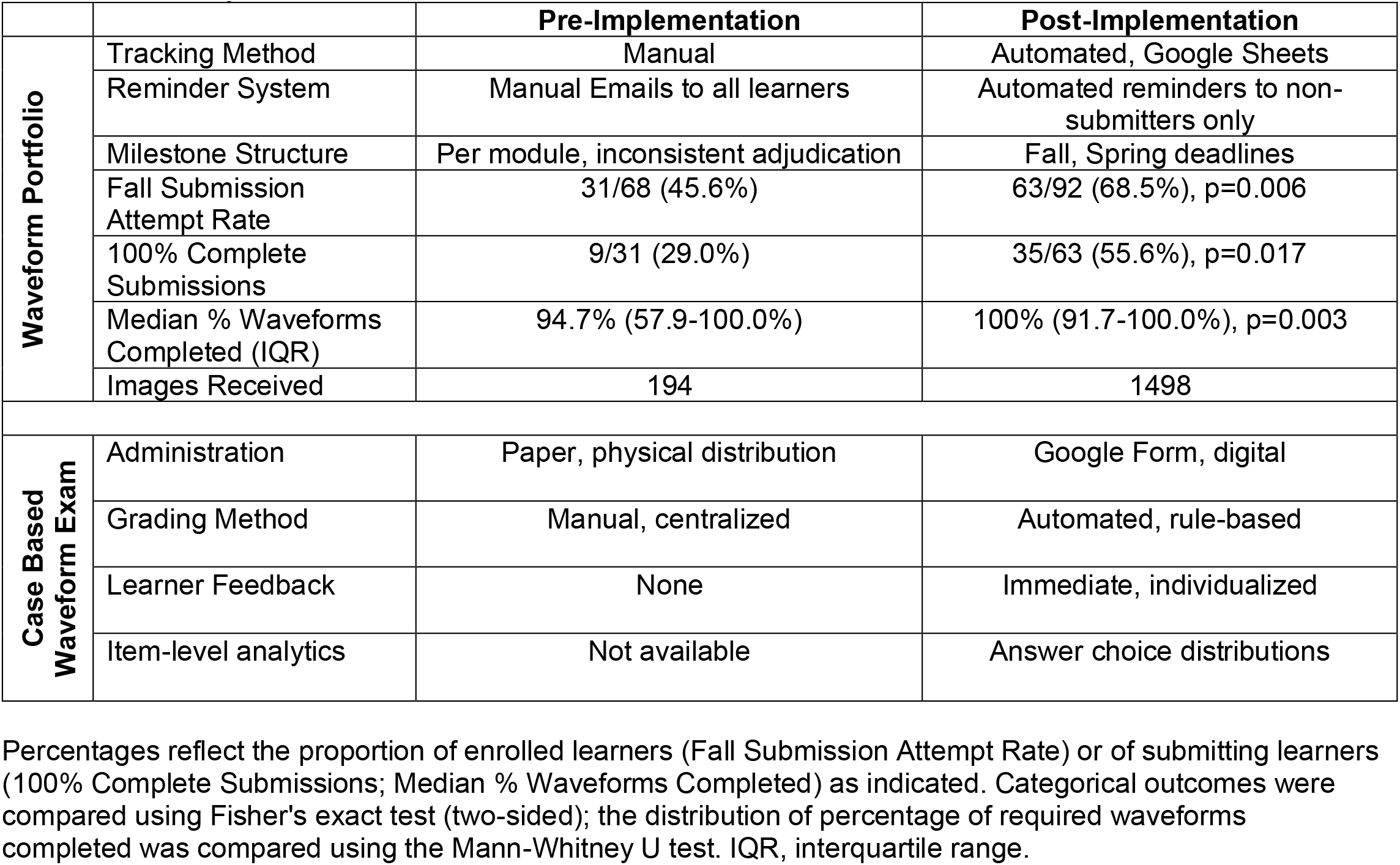
Portfolio Completion and Examination Grading Outcomes Before and After Automated Digital Infrastructure Implementation.

|  |  | **Pre-Implementation** | **Post-Implementation** |
| --- | --- | --- | --- |
| **Waveform Portfolio** | Tracking Method | Manual | Automated, Google Sheets |
|  | Reminder System | Manual Emails to all learners | Automated reminders to non-submitters only |
|  | Milestone Structure | Per module, inconsistent adjudication | Fall, Spring deadlines |
|  | Fall Submission Attempt Rate | 31/68 (45.6%) | 63/92 (68.5%), p=0.006 |
|  | 100% Complete Submissions | 9/31 (29.0%) | 35/63 (55.6%), p=0.017 |
|  | Median % Waveforms Completed (IQR) | 94.7% (57.9-100.0%) | 100% (91.7-100.0%), p=0.003 |
|  | Images Received | 194 | 1498 |
| **Case Based Waveform Exam** | Administration | Paper, physical distribution | Google Form, digital |
|  | Grading Method | Manual, centralized | Automated, rule-based |
|  | Learner Feedback | None | Immediate, individualized |
|  | Item-level analytics | Not available | Answer choice distributions |

Percentages reflect the proportion of enrolled learners (Fall Submission Attempt Rate) or of submitting learners (100% Complete Submissions; Median % Waveforms Completed) as indicated. Categorical outcomes were compared using Fisher's exact test (two-sided); the distribution of percentage of required waveforms completed was compared using the Mann-Whitney U test. IQR, interquartile range.

### Manual Examination Grading Workload

Manual grading of waveform examinations required a mean of 5.1 minutes per examination (Standard Deviation (SD) 0.59 minutes). For a 100-learner cohort, this projects to approximately 8.5 grading-hours per faculty grader (100 tests × 5.1 minutes ÷ 60 minutes/hour) and SD of 1.0 hours.

### Automated Examination Grading Analytics

Retrospective processing of previously administered examinations through the automated grading system confirmed accurate operationalization of the expert-defined scoring rubric. The system generated item-level answer choice distributions for each examination question, enabling identification of frequently selected distractors and patterns of learner misunderstanding not readily apparent through manual review.

## Discussion

Standardizing waveform portfolio requirements and implementing automated milestone tracking improved timely formative assessment completion, while a rule-based automated grading system enabled scalable summative assessment with individualized learner feedback and item-level analytic outputs. Together, these interventions address the parallel administrative and content scarcity constraints that had limited a large-scale mechanical ventilation education program’s ability to scale without proportional increases in faculty burden.

Improvements in portfolio submission rates and completion score distributions suggest more thorough and consistent engagement. The proportion of learners completing all required waveforms nearly doubled, and the completion IQR itself shifted upward and narrowed (57.9-100.0% to 91.7-100%). No formal consequences for incomplete submissions were imposed, suggesting the observed shift reflects engagement rather than compliance pressure. Fall 2025 submission requirements included 24 required waveforms compared with 19 in Fall 2024, meaning improved submission rates were also achieved against a more demanding standard. Overall submission rates leave room for continued improvement, and future work should examine barriers to participation among non-submitters, including clinical workload and competing demands across sites. Automated grading is projected to eliminate 8.5 faculty hours per grader per 100 tests. This conservative estimate reflects grading time alone and does not account for the additional burden of score tabulation, intergrader reconciliation, feedback generation, and item-level analysis. The first phase, encompassing registration and program orientation, represents a remaining opportunity for future work. The shift from manual to automated administrative workflows reflects a deliberate reallocation of expert time toward higher-value activities. In a one-on-one preceptorial model sustained entirely by volunteer expert educators and learners, the irreplaceable core is the teaching relationship, not portfolio tracking, reminder emails, or examination grading.

Imposing financial costs through licensing fees, proprietary platforms, or IT procurement would further burden volunteers from multiple institutions whose contribution is already measured in time and expertise. Academic medicine has long grappled with the erosion of protected teaching time under mounting clinical productivity demands[16], a pressure that has only intensified in recent years. By building on zero-cost, universally accessible tools requiring no institutional infrastructure, these systems honor the commitment of expert educators and learners who sustain the program by ensuring that financial burden is never added to the time and expertise they already contribute.

Automation also closed a feedback loop previously structurally absent from the program. Paper-based administration and centralized manual grading previously precluded timely, individualized feedback since learners completed a summative assessment without receiving meaningful information about their performance. Automated grading enables rapid return of individualized results, providing an opportunity for directed learning (Supplemental Figure 3a). Item-level analytics will allow identification of frequently selected distractors and patterns of misunderstanding, informing iterative refinement of examination content and curriculum design (Supplemental Figure 3b).

The approach described here has broad implications beyond mechanical ventilation. Subspecialty domains such as point-of-care ultrasound, ICU physiology, and procedural skill development face analogous challenges, frequent formative assessment requirements, limited expert pools, and growing cohorts that outpace traditional infrastructure. The infrastructure described here demonstrates that zero-cost tools built outside proprietary ecosystems can meet the scalability demands of competency-based training, offering a transferable model for programs navigating similar constraints.

The WP system also has the potential to address the content scarcity that has constrained frequent assessment deployment. Learner-generated portfolios, adjudicated by expert faculty, represent a self-reinforcing pipeline for future examination content with each cohort’s submissions enriching the case library available for subsequent cycles. The increased volume of waveform submissions following standardization also highlights an important distinction inherent to automated tracking systems and future opportunities for study. The platform facilitated measurement of submission completeness and timeliness rather than clinical accuracy of submitted content. Digital infrastructure can manage the logistics of longitudinal assessment, but the interpretive judgment required to validate clinically complex submissions cannot be automated. This distinction is critical: the goal of these systems is to protect expert time for expert work, not to replace it.

### Limitations

The pre-post design without randomization limits causal inference. Although standardization of portfolio requirements was implemented as a deliberate first step prior to automation, the study design did not include an intermediate assessment of portfolio completion rates following standardization alone, so relative contributions of standardization and automation cannot be disentangled. Concurrent changes between cohorts (i.e. growth in enrollment, number of participating sites, and program maturity) may have independently contributed to observed improvement in portfolio submission rates. This study evaluated timely portfolio submission rather than direct measures of sustained knowledge retention or clinical performance. Future studies should examine whether improved portfolio completion translates into measurable differences in ventilator management competency.

Prospective deployment of CBWE grading system beginning in Summer 2026 will provide the first real-world assessment of operational impact on faculty workload, learner experience, and integration of examination analytics into curriculum improvement processes.

Learner-generated WPs represent a self-reinforcing pipeline for future examination content, creating a virtuous cycle between formative and summative assessment that strengthens the program’s long-term sustainability. By reallocating administrative burden away from expert faculty, this infrastructure preserves the high-fidelity teaching relationship at the core of the preceptorial model while enabling it to operate on a national scale. By protecting expert time for expert work, this infrastructure ensures that the preceptorial model, built on the irreplaceable teaching relationship between expert and learner, can grow without sacrificing the fidelity that defines it.

## Data Availability

All data produced in the present study are available upon reasonable request to the authors.

---

**Supplementary Figure 1.**
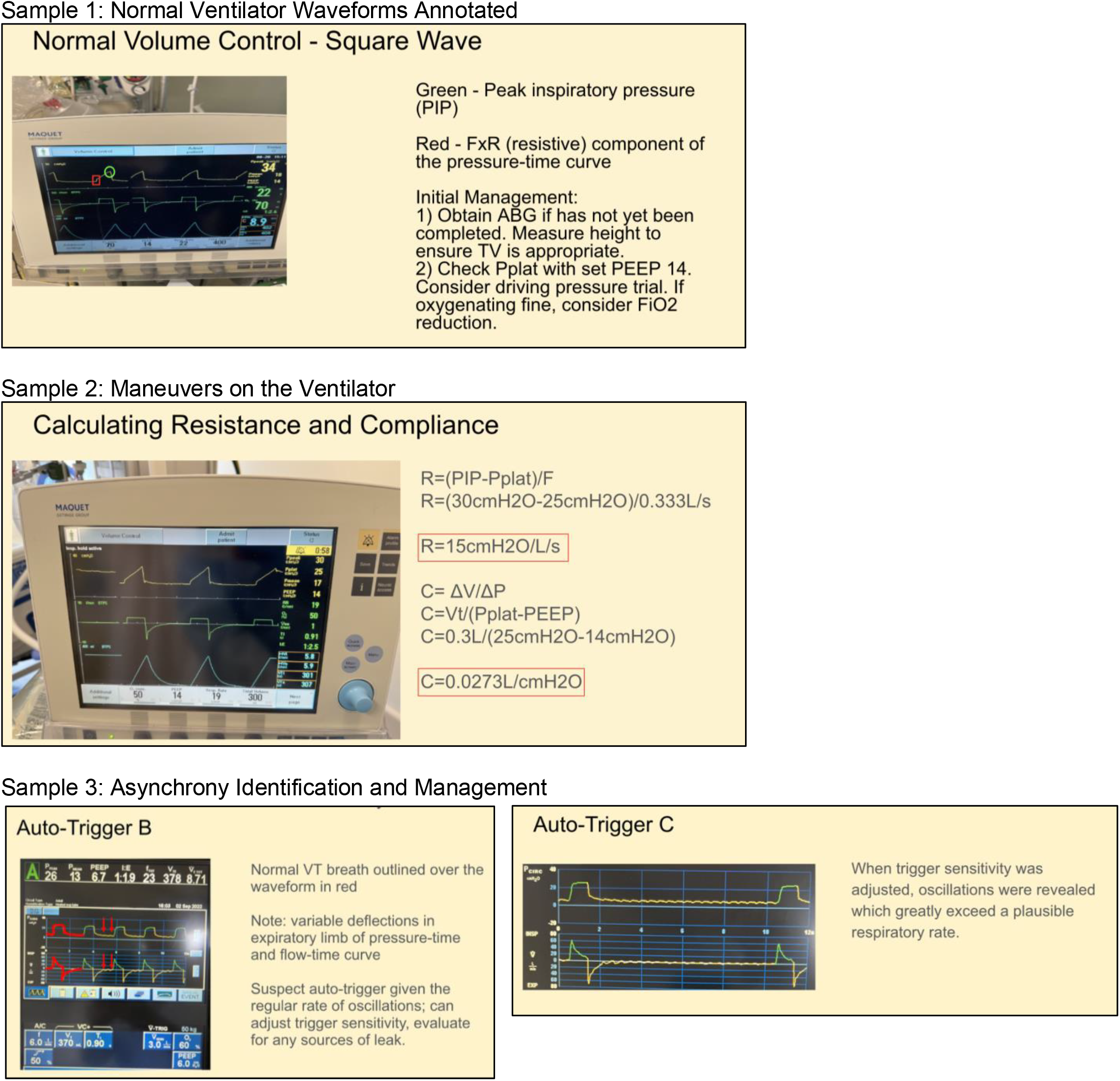
Sample Waveform Portfolio Submissions. Representative examples of structured submission template (waveform image, physiologic interpretation, and clinical management documentation) applied to a normal waveform, a maneuver, and an asynchrony.

**Supplementary Figure 2.**
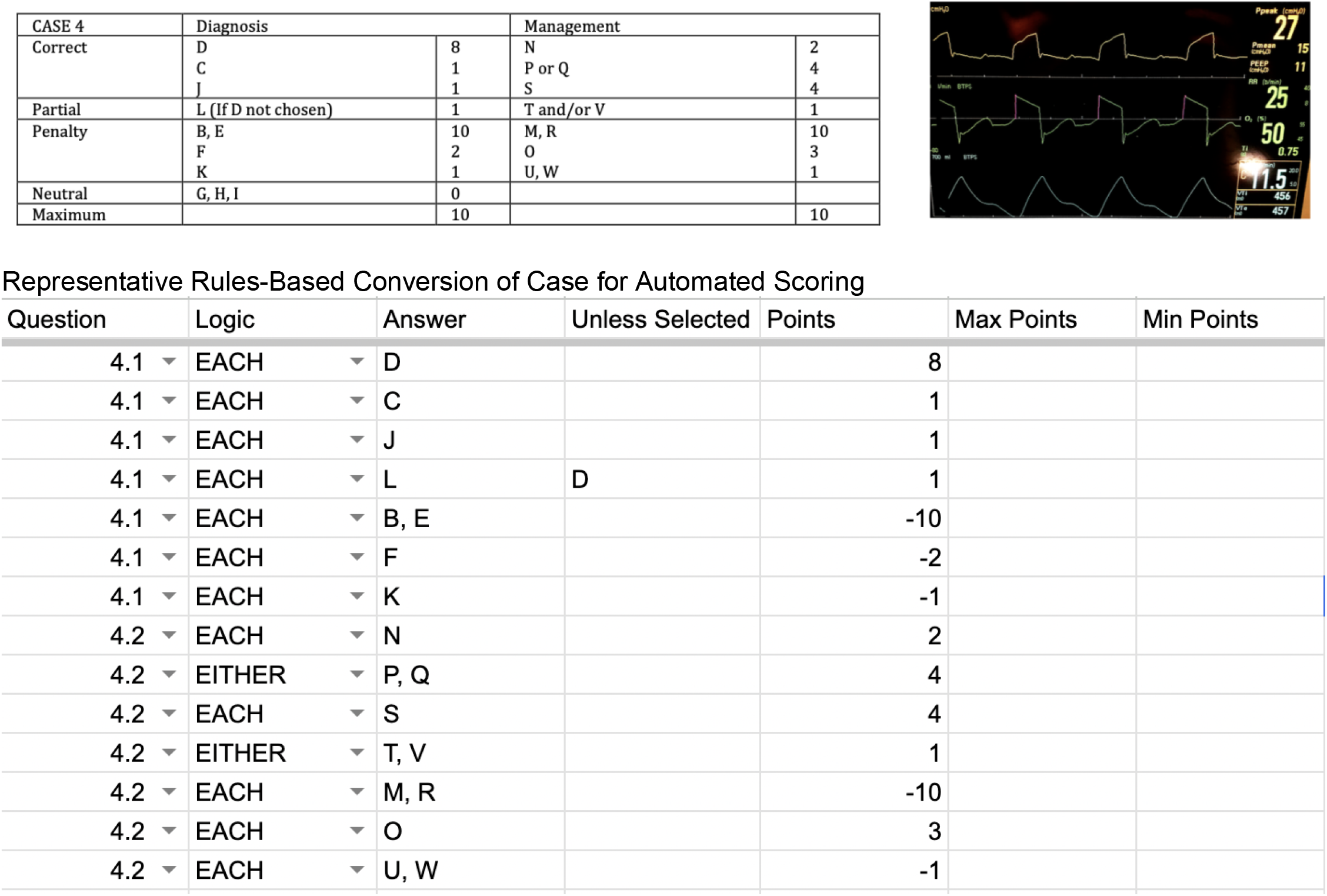
Representative Case Rubric from Written CBWE^a^ ^a^Note: Letter codes represent answer choices to protect exam integrity while demonstrating rubric structure.

**Supplementary Figure 3a.**
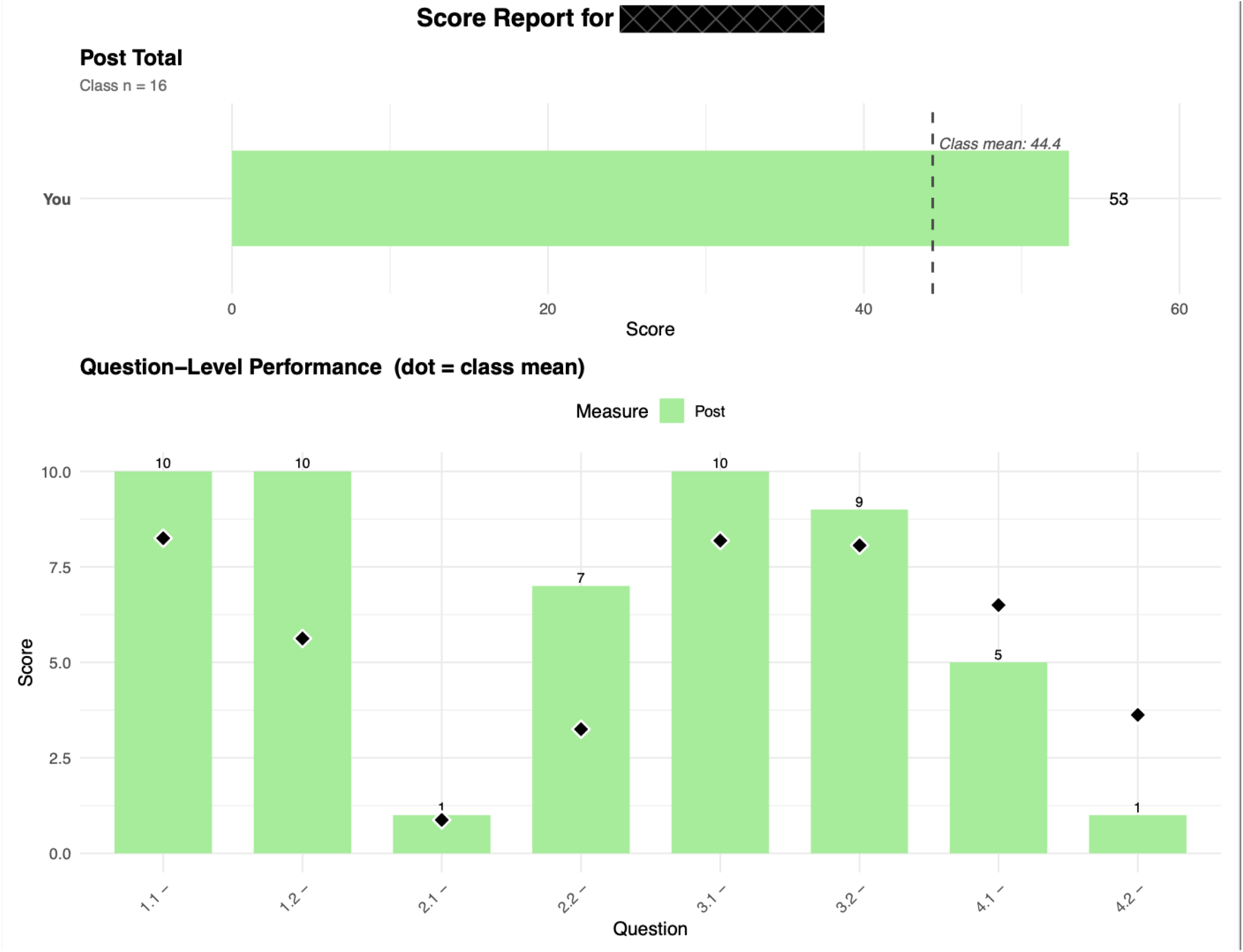
Sample Deidentified Learner Score Report

**Supplementary Figure 3b.**
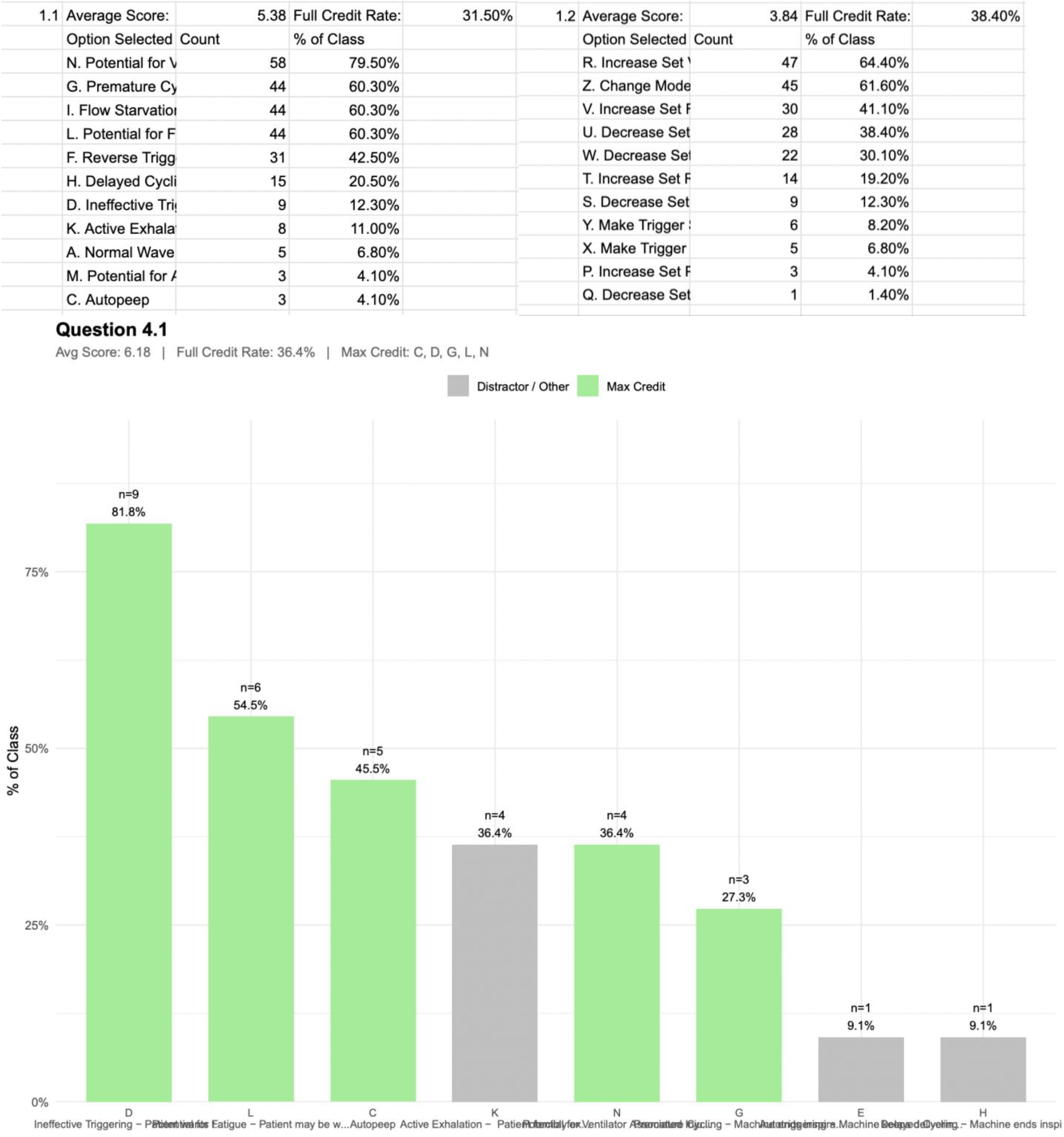
Example Answer-Choice Automated Analytics for Single Case with Educator Summary Review Sheet

## Notes

### Competing Interest Statement

The authors have declared no competing interest.

### Author Declarations

IRB of National Institutes of Health Clinical Center deemed this educational research project exempt

