## Supplementary material for "Zero-Cost Digital Automation for Scalable Assessment in a National Mechanical Ventilation Curriculum": Code

**Supplemental Information – Raw Code**

**Learner Reminder Email Code**

/***************************************************************

* LEARNER REMINDERS — Waveform Portfolio Tracker

* Sends weekly emails to learners who have not "Submitted".

* Depends on config.gs for SHEET_LEARNERS, getSS_(), parseEmails_(),

* and WEEKLY_END_DATE_YYYYMMDD (auto-stop date).

***************************************************************/

// ===== Settings to tweak =====

const REMINDERS_LOG_SHEET = 'Reminders'; // audit log sheet name

const REMINDER_SUBJECT = 'Friendly reminder: Please submit your waveform portfolio';

const REMINDER_COOLDOWN_DAYS = 7; // don’t email a learner more often than this

const REMINDER_BCC = ''; // e.g. '' or leave ''

const REMINDER_REPLY_TO = ''; // optional reply-to for replies

// Simple template supports {{NAME}} and {{SHEET_URL}}

const REMINDER_BODY_HTML =

'Hi {{NAME}},<br><br>' +

'This is a friendly reminder to submit your Fall 2025 waveform portfolio with any edits/missing images for the <b>Mechanical Ventilator Preceptorial</b>.' +

'Best,<br>MV Preceptorial Team<br>' +

'<i>(This is an automated reminder, please contact the team directly with any questions.)</i>';

// ========== Public functions (show up in Run/Triggers) ==========

/**

* Sends reminders to all learners whose "Submitted" (col C) is not TRUE.

* Respects a per-learner cooldown (REMINDER_COOLDOWN_DAYS).

* Auto-stops after WEEKLY_END_DATE_YYYYMMDD.

*/

function sendPortfolioReminders() {

if (isPastReminderEndDate_()) {

disablePortfolioReminders_();

console.log('Reminders disabled — past end date.');

return;

}

const ss = getSS_();

const learners = ss.getSheetByName(SHEET_LEARNERS);

if (!learners) { console.log('Learners sheet not found.'); return; }

const log = ensureRemindersLog_();

const sheetUrl = ss.getUrl();

const now = new Date();

const cooldownMs = REMINDER_COOLDOWN_DAYS * 24 * 60 * 60 * 1000;

// Read Learners: Name(A), Email(B), Submitted(C), Submitted At(D), File URL(E)

const last = learners.getLastRow();

if (last < 2) { console.log('No learners.'); return; }

const rows = learners.getRange(2, 1, last - 1, 5).getValues();

// Build per-row "last reminded" map from the log

const lastRemindedByRow = buildLastRemindedIndex_(log);

let sent = 0, skippedSubmitted = 0, skippedNoEmail = 0, skippedCooldown = 0;

for (let i = 0; i < rows.length; i++) {

const rowNum = i + 2;

const name = String(rows[i][0] || '').trim();

const emailCell = String(rows[i][1] || '').trim();

const submitted = rows[i][2] === true || String(rows[i][2]).toUpperCase() === 'TRUE';

if (submitted) { skippedSubmitted++; continue; }

const emails = parseEmails_(emailCell); // from config.gs

if (!emails.length) { skippedNoEmail++; continue; }

// Cooldown check

const lastRem = lastRemindedByRow.get(rowNum);

if (lastRem && (now.getTime() - lastRem.getTime()) < cooldownMs) {

skippedCooldown++;

continue;

}

// Personalize and send

const firstName = extractFirstName_(name);

const htmlBody = formatTemplate_(REMINDER_BODY_HTML, {

NAME: firstName || 'there',

SHEET_URL: sheetUrl

});

const mail = { to: emails.join(','), subject: REMINDER_SUBJECT, htmlBody };

if (REMINDER_BCC) mail.bcc = REMINDER_BCC;

if (REMINDER_REPLY_TO) mail.replyTo = REMINDER_REPLY_TO;

MailApp.sendEmail(mail);

sent++;

appendReminderLog_(log, now, rowNum, name, emails.join(','));

Utilities.sleep(50); // gentle throttle

}

console.log(

'Reminder run complete — sent=' + sent +

', alreadySubmitted=' + skippedSubmitted +

', noEmail=' + skippedNoEmail +

', cooldown=' + skippedCooldown

);

}

/**

* One-time helper: create a weekly trigger (e.g., Mondays at 9am).

*/

function createLearnerReminderTrigger() {

ScriptApp.newTrigger('sendPortfolioReminders')

.timeBased()

.everyWeeks(1)

.onWeekDay(ScriptApp.WeekDay.MONDAY)

.atHour(9)

.create();

console.log('Learner reminder trigger created: Mondays 09:00.');

}

/**

* Optional: send a single preview to yourself.

*/

function sendPortfolioRemindersPreview() {

const ss = getSS_();

const sheetUrl = ss.getUrl();

const htmlBody = formatTemplate_(REMINDER_BODY_HTML, {

NAME: 'Learner Name',

SHEET_URL: sheetUrl

});

MailApp.sendEmail({

to: Session.getActiveUser().getEmail(),

subject: '[Preview] ' + REMINDER_SUBJECT,

htmlBody

});

console.log('Preview sent to your email.');

}

// ========== Internal helpers (kept “private”) ==========

// Stop reminders after the configured end date in config.gs

function isPastReminderEndDate_() {

// Re-use the same end date constant used for weekly summaries

const tz = Session.getScriptTimeZone();

const today = Utilities.formatDate(new Date(), tz, 'yyyy-MM-dd');

return (typeof WEEKLY_END_DATE_YYYYMMDD === 'string') && today > WEEKLY_END_DATE_YYYYMMDD;

}

// Delete triggers for this handler so it stops after the end date

function disablePortfolioReminders_() {

ScriptApp.getProjectTriggers().forEach(t => {

if (t.getHandlerFunction && t.getHandlerFunction() === 'sendPortfolioReminders') {

ScriptApp.deleteTrigger(t);

}

});

PropertiesService.getScriptProperties().setProperty('LEARNER_REMINDERS_PAUSED', '1');

}

function ensureRemindersLog_() {

const ss = getSS_();

let log = ss.getSheetByName(REMINDERS_LOG_SHEET);

if (!log) {

log = ss.insertSheet(REMINDERS_LOG_SHEET);

log.getRange(1, 1, 1, 5).setValues([[

'Timestamp', 'Learner Row', 'Name', 'Emails', 'Note'

]]);

}

return log;

}

function appendReminderLog_(logSheet, when, rowNum, name, emails) {

logSheet.appendRow([ when, rowNum, name, emails, 'sent' ]);

}

function buildLastRemindedIndex_(logSheet) {

const last = logSheet.getLastRow();

const m = new Map();

if (last < 2) return m;

const data = logSheet.getRange(2, 1, last - 1, 5).getValues();

for (let i = 0; i < data.length; i++) {

const when = data[i][0];

const rowNum = data[i][1];

if (!rowNum || !when) continue;

const prev = m.get(rowNum);

const curr = new Date(when);

if (!prev || curr > prev) m.set(rowNum, curr);

}

return m;

}

// Tiny mustache-like replacer for {{KEY}}

function formatTemplate_(tpl, data) {

return tpl.replace(/\{\{(\w+)\}\}/g, function(_, k){

return (k in data) ? String(data[k]) : '';

});

}

**Weekly Email Blast Code**

/***************************************************************

* WEEKLY EMAIL SUMMARY - AI in MedEd: MV Preceptorial Tracker

* Requires: config.gs + tracker.gs

* Uses constants: WEEKLY_RECIPIENTS, WEEKLY_END_DATE_YYYYMMDD

***************************************************************/

// Check if we're past stop date

function isPastEndDate_() {

const tz = Session.getScriptTimeZone();

const today = Utilities.formatDate(new Date(), tz, 'yyyy-MM-dd');

return today > WEEKLY_END_DATE_YYYYMMDD;

}

// Disable future triggers once end date passes

function disableWeeklySummary_() {

ScriptApp.getProjectTriggers().forEach(t => {

if (t.getHandlerFunction && t.getHandlerFunction() === 'sendWeeklySummary') {

ScriptApp.deleteTrigger(t);

}

});

PropertiesService.getScriptProperties().setProperty('WEEKLY_SUMMARY_PAUSED', '1');

Logger.log('Weekly summary disabled after end date.');

}

/**

* Main weekly summary sender

* Always sends a weekly email, even if no new submissions

*/

function sendWeeklySummary() {

// Stop sending after configured date

if (isPastEndDate_()) {

disableWeeklySummary_();

console.log('Weekly summary skipped — past end date.');

return;

}

const ss = getSS_();

const subs = ss.getSheetByName(SHEET_SUBMISSIONS);

const learners = ss.getSheetByName(SHEET_LEARNERS);

if (!subs || !learners) {

console.log('Missing Learners or Submissions sheet.');

return;

}

const props = PropertiesService.getScriptProperties();

const LAST_WEEKLY_KEY = 'LAST_WEEKLY_ISO';

const now = new Date();

const oneWeekAgo = new Date(now.getTime() - 7 * 24 * 60 * 60 * 1000);

const lastDigestISO = props.getProperty(LAST_WEEKLY_KEY);

const windowStart = lastDigestISO

? new Date(Math.max(oneWeekAgo.getTime(), new Date(lastDigestISO).getTime()))

: oneWeekAgo;

// Collect new submissions

const lastRow = subs.getLastRow();

const data = lastRow > 1 ? subs.getRange(2, 1, lastRow - 1, 8).getValues() : [];

const newRows = data.filter(r => r[0] && new Date(r[0]) > windowStart);

// Compute overall progress

const learnerRows = learners.getLastRow() > 1

? learners.getRange(2, 1, learners.getLastRow() - 1, 5).getValues()

: [];

const totalLearners = learnerRows.length;

const submittedCount = learnerRows.filter(r => r[2] === true || String(r[2]).toUpperCase() === 'TRUE').length;

const submissionRate = totalLearners ? ((submittedCount / totalLearners) * 100).toFixed(1) + '%' : 'N/A';

const sheetUrl = ss.getUrl();

const total = newRows.length;

const matched = newRows.filter(r => r[6] && String(r[6]).trim()).length;

const unmatched = total - matched;

// Compose message

const subject = `📬 Weekly Upload Summary — ${ss.getName()} (${total} new)`;

const headerLine = 'Hi Everyone! Here is the upload summary for the past week for the Mechanical Ventilator Preceptorial.';

const header = ['Date', 'File', 'Learner (matched?)', 'Owner'];

const recentRows = newRows.slice(-20).map(r => [

formatDate_(r[0]),

truncate_(r[1], 45),

r[6] ? r[6] + ' ✓' : '(no match)',

r[4] || r[5] || ''

]);

const body =

`<b>${headerLine}</b><br><br>` +

`📊 <b>Weekly Upload Summary</b><br><br>` +

`<b>Spreadsheet:</b> <a href="${sheetUrl}">${ss.getName()}</a><br><br>` +

`<b>Time window:</b> ${formatDate_(windowStart)} → ${formatDate_(now)}<br>` +

`<b>New submissions:</b> ${total}<br>` +

`<b>Matched learners:</b> ${matched}<br>` +

`<b>Unmatched:</b> ${unmatched}<br><br>` +

`<b>Total learners:</b> ${totalLearners}<br>` +

`<b>Submitted learners:</b> ${submittedCount}<br>` +

`<b>Submission rate:</b> ${submissionRate}<br><br>` +

`Recent (up to 20 shown):<br>` +

`<pre>${asFixedWidthTable_([header].concat(recentRows))}</pre><br>` +

`Click the link above to view the full tracker spreadsheet.<br><br>` +

`<i>Automatic summary generated on ${formatDate_(now)}</i>`;

const recipients = WEEKLY_RECIPIENTS.join(',');

MailApp.sendEmail({

to: recipients,

subject: subject,

htmlBody: body

});

props.setProperty(LAST_WEEKLY_KEY, now.toISOString());

console.log(`Weekly summary sent to ${recipients}. ${total} new rows.`);

}

/**

* One-time setup: create a weekly trigger (e.g. Mondays at 8am)

*/

function createWeeklyTrigger() {

ScriptApp.newTrigger('sendWeeklySummary')

.timeBased()

.everyWeeks(1)

.onWeekDay(ScriptApp.WeekDay.MONDAY)

.atHour(8) // 24h format, project timezone

.create();

console.log('Weekly summary trigger created.');

}

**Tracker Code**

/**** TRACKER: scan + match + mark (uses config.gs) ****/

function scanFolderTree() {

// From config.gs

const { learnersSh, submissionsSh } = ensureTabs_();

const props = PropertiesService.getScriptProperties();

const processedPrefix = 'PROCESSED_';

const lastRunKey = 'LAST_RUN_ISO';

const lastRunIso = props.getProperty(lastRunKey);

const lastRun = lastRunIso ? new Date(lastRunIso) : null;

// Roster + indexes

const roster = loadRoster_(learnersSh);

const nameIndex = buildNameIndex_(roster);

const emailIndex = buildEmailIndex_(roster);

const root = DriveApp.getFolderById(FOLDER_ID); // from config.gs

const now = new Date();

let processedCount = 0;

walkFolderTree_(root, function(file) {

if (file.isTrashed && file.isTrashed()) return;

const fileId = file.getId();

if (props.getProperty(processedPrefix + fileId)) return; // already handled

// Advanced Drive (enable "Drive API" in Services)

const meta = Drive.Files.get(fileId, {

fields: 'id,name,webViewLink,owners(emailAddress),lastModifyingUser(emailAddress),modifiedTime,trashed'

});

if (meta.trashed) return;

// Skip unchanged since last run

if (lastRun && meta.modifiedTime && new Date(meta.modifiedTime) <= lastRun) return;

const fileName = meta.name || file.getName();

const fileUrl = meta.webViewLink || file.getUrl();

const ownerEmail = (meta.owners && meta.owners[0] && meta.owners[0].emailAddress) ? meta.owners[0].emailAddress : '';

const lastModEmail = (meta.lastModifyingUser && meta.lastModifyingUser.emailAddress) ? meta.lastModifyingUser.emailAddress : '';

// Match learner (email → filename)

let match = null, method = '';

if (lastModEmail && emailIndex.has(lastModEmail.toLowerCase())) {

match = emailIndex.get(lastModEmail.toLowerCase()); method = 'email:lastModifyingUser';

} else if (ownerEmail && emailIndex.has(ownerEmail.toLowerCase())) {

match = emailIndex.get(ownerEmail.toLowerCase()); method = 'email:owner';

} else {

const byName = matchByFilename_(fileName, nameIndex);

if (byName) {

// Was it an initials pattern like "doe j" / "doe, j" / "doe-j" / "[doe, j]"?

const parts = splitNameParts_(byName.name);

const ln = parts.last, fi = parts.first ? parts.first.charAt(0) : '';

const f = normalizeText_(fileName);

const initialsHit =

(ln && fi) && (

new RegExp('\\b'+ln+'\\s+'+fi+'\\b').test(f) ||

new RegExp('\\b'+ln+'\\s*,\\s*'+fi+'\\b').test(f) ||

new RegExp('\\b'+ln+'[_-]'+fi+'\\b').test(f) ||

new RegExp('\\['+'\\s*'+ln+'\\s*,\\s*'+fi+'\\s*\\]').test(f)

);

match = byName;

method = initialsHit ? 'filename:initial' : 'filename:name';

}

}

// Log + mark submission

submissionsSh.appendRow([

now, fileName, fileUrl, fileId,

ownerEmail, lastModEmail,

match ? match.name : '', method

]);

if (match) markSubmission_(learnersSh, match.row, fileUrl, now);

props.setProperty(processedPrefix + fileId, '1');

processedCount++;

Utilities.sleep(35);

});

props.setProperty(lastRunKey, now.toISOString());

console.log('Processed ' + processedCount + ' files at ' + now.toISOString());

}

// Depth-first traversal over folders

function walkFolderTree_(folder, onFile) {

const files = folder.getFiles();

while (files.hasNext()) onFile(files.next());

const subs = folder.getFolders();

while (subs.hasNext()) walkFolderTree_(subs.next(), onFile);

}

/*************** Roster + matching helpers ****************/

function loadRoster_(sheet) {

const last = sheet.getLastRow();

if (last < 2) return [];

const rng = sheet.getRange(2,1,last-1,5).getValues();

const out = [];

for (let i=0;i<rng.length;i++){

const r = rng[i];

out.push({

row: i+2,

name: (typeof r[0] === 'string' ? r[0].trim() : ''),

email:(typeof r[1] === 'string' ? r[1].trim() : ''),

submitted: r[2],

submittedAt: r[3],

fileUrl: r[4]

});

}

return out;

}

function buildEmailIndex_(roster){

const m = new Map();

roster.forEach(function(r){

if (!r.email) return;

parseEmails_(r.email).forEach(function(e){

m.set(e, r); // index each email → same learner record

});

});

return m;

}

function buildNameIndex_(roster){

const m = new Map();

roster.forEach(function(r){

if (!r.name) return;

const norm = normalizeName_(r.name);

const flipped = flipName_(norm);

m.set(norm, r);

if (flipped) m.set(flipped, r);

});

return m;

}

function splitNameParts_(fullName){

const t = normalizeName_(fullName);

const parts = t.split(/\s+/).filter(Boolean);

if (parts.length === 0) return {first:'', last:''};

if (parts.length === 1) return {first:parts[0], last:''};

return {first:parts[0], last:parts[parts.length-1]};

}

function matchByFilename_(fileName, nameIndex){

const f = normalizeText_(fileName);

// 1) full-name / variant

for (const [variant, record] of nameIndex.entries()){

if (!variant) continue;

const re = new RegExp('\\b' + escapeRegex_(variant) + '\\b');

if (re.test(f)) return record;

}

// 2) bigrams

const tokens = f.split(/\s+/).filter(Boolean);

for (let i=0;i<tokens.length-1;i++){

const bigram = tokens[i] + ' ' + tokens[i+1];

if (nameIndex.has(bigram)) return nameIndex.get(bigram);

}

// (3) initials handled in scanFolderTree when a name hit is chosen

return null;

}

// Accept comma/semicolon/space-separated emails in one cell

function parseEmails_(cell){

return String(cell || '')

.split(/[,;\s]+/)

.map(s => s.trim().toLowerCase())

.filter(Boolean);

}

// Mark "Submitted" in Learners (C) and set date/url (D/E)

function markSubmission_(sheet, row, fileUrl, when){

const SUBMITTED_COL = 3, AT_COL = 4, URL_COL = 5;

const vals = sheet.getRange(row,1,1,5).getValues()[0];

const already = vals[SUBMITTED_COL-1] === true || String(vals[SUBMITTED_COL-1]).toUpperCase() === 'TRUE';

if (!already){

sheet.getRange(row, SUBMITTED_COL).setValue(true);

sheet.getRange(row, AT_COL).setValue(when);

sheet.getRange(row, URL_COL).setValue(fileUrl);

} else if (!vals[URL_COL-1]) {

sheet.getRange(row, URL_COL).setValue(fileUrl);

}

}

/*************** Debug helpers (optional) ****************/

function debugTreeSummary() {

const root = DriveApp.getFolderById(FOLDER_ID);

let folders = 0, files = 0;

(function walk(f) {

const fs = f.getFiles();

while (fs.hasNext()) { fs.next(); files++; if (files >= 5000) break; }

const subs = f.getFolders();

while (subs.hasNext()) { folders++; walk(subs.next()); if (files >= 5000) break; }

})(root);

Logger.log('Found ~' + folders + ' folders and ~' + files + ' files under root.');

}

function clearLastRun() {

PropertiesService.getScriptProperties().deleteProperty('LAST_RUN_ISO');

Logger.log('LAST_RUN_ISO cleared');

}

**Score Report Generator**

\# ----------------------------

> # 1) CSV File Read

> # ----------------------------

read_tracker <- function(path, id_col = "Learner") {

+	for (skip_n in 0:3) {

+	df <- tryCatch(

+	readr::read_csv(path, skip = skip_n, show_col_types = FALSE),

+	error = function(e) NULL

+	)

+

+	if (!is.null(df)) {

+	names(df) <- trimws(names(df))

+	if (id_col %in% names(df)) {

+	message("Read file successfully using skip = ", skip_n)

+	return(df)

+	}

+	}

+	}

+

+	stop(

+	paste0(

+	"Could not find column '", id_col, "' after trying skip = 0, 1, 2, and 3.\n",

+	"Please confirm the learner column is named exactly '", id_col, "'."

+	)

+	)

+ }

> > tracker <- read_tracker(input_file, id_col = id_col)

\# ----------------------------

> # 2) Clean column names

> # ----------------------------

> names(tracker) <- trimws(names(tracker))

> > # Remove junk unnamed columns like ...1, ...28

> bad_cols <- grepl("^\\.\\.\\.[0-9]+$", names(tracker))

> if (any(bad_cols)) {

+	message("Removing unnamed columns: ", paste(names(tracker)[bad_cols], collapse = ", "))

+	tracker <- tracker[, !bad_cols, drop = FALSE]

> > # Ensure Email column exists

> if (!email_col %in% names(tracker)) {

+	tracker[[email_col]] <- NA_character_

+ }

> > # Try to locate OTHER column flexibly if exact name not found

> if (!other_col %in% names(tracker)) {

+	other_matches <- names(tracker)[toupper(names(tracker)) == "OTHER"]

+	if (length(other_matches) > 0) {

+	other_col <- other_matches[1]

+	}

+ }

> > # ----------------------------

> # 3) Clean learner rows

> # ----------------------------

> tracker <- tracker %>%

+	mutate(across(all_of(c(id_col, email_col)), as.character)) %>%

+	mutate(across(all_of(c(id_col, email_col)), ~ trimws(.))) %>%

+	filter(!is.na(.data[[id_col]]), .data[[id_col]] != "")

> > if (nrow(tracker) == 0) {

+	stop("No learner rows found after cleaning.")

+ }

> > # ----------------------------

> # 4) Split fixed items vs OTHER

> # ----------------------------

> all_non_id_cols <- setdiff(names(tracker), c(id_col, email_col))

> > fixed_item_cols <- setdiff(all_non_id_cols, other_col)

> > if (length(fixed_item_cols) == 0) {

+	stop("No fixed waveform item columns were found.")

+ }

> > # Convert fixed items to character for safe pivoting

> tracker <- tracker %>%

+	mutate(across(all_of(fixed_item_cols), as.character))

> > # ----------------------------

> # 5) Helper functions

> # ----------------------------

> normalize_status <- function(x) {

+	x_chr <- trimws(as.character(x))

+

+	dplyr::case_when(

+	is.na(x_chr) ~ "missing",

+	x_chr == "1" ~ "complete",

+	toupper(x_chr) == "T" ~ "text_only",

+	x_chr == "0" ~ "missing",

+	x_chr == "" ~ "missing",

+	TRUE ~ "missing"

+	)

+ }

> > get_section <- function(item_name, section_map) {

+	for (sec in names(section_map)) {

+	if (item_name %in% section_map[[sec]]) return(sec)

+	}

+	return("Other")

+ }

> > safe_filename <- function(x) {

+	x <- as.character(x)

+	x <- str_replace_all(x, "[^A-Za-z0-9]+", "_")

+	x <- str_replace_all(x, "^_|_$", "")

+	ifelse(x == "", "learner", x)

+ }

> > collapse_or_blank <- function(x) {

+	x <- x[!is.na(x)]

+	if (length(x) == 0) "" else paste(x, collapse = "; ")

+ }

> > # ----------------------------

> # 6) Long table for fixed items

> # ----------------------------

> tracker_long <- tracker %>%

+	pivot_longer(

+	cols = all_of(fixed_item_cols),

+	names_to = "item",

+	values_to = "raw_value"

+	) %>%

+	mutate(

+	status = normalize_status(raw_value),

+	section = vapply(item, get_section, character(1), section_map = section_map),

+	display_item = ifelse(item %in% names(name_map), name_map[item], item)

+	)

> > # ----------------------------

> # 7) Summary for fixed items

> # ----------------------------

> fixed_summary <- tracker_long %>%

+	group_by(.data[[id_col]], .data[[email_col]]) %>%

+	summarise(

+	n_complete = sum(status == "complete", na.rm = TRUE),

+	n_text_only = sum(status == "text_only", na.rm = TRUE),

+	n_missing = sum(status == "missing", na.rm = TRUE),

+	n_total_fixed = dplyr::n(),

+	pct_complete_fixed = round(100 * n_complete / n_total_fixed, 1),

+	.groups = "drop"

+	)

> > # ----------------------------

> # 8) Section-level grouped details

> # ----------------------------

> grouped_details <- tracker_long %>%

+	filter(status != "complete") %>%

+	group_by(.data[[id_col]], section, status) %>%

+	summarise(

+	items = list(display_item),

+	.groups = "drop"

+	)

> > # ----------------------------

> # 9) OTHER requirement handling

> #	One numeric column; learners need 2 varied waveforms

> # ----------------------------

> if (other_col %in% names(tracker)) {

+	other_df <- tracker %>%

+	transmute(

+	!!id_col := .data[[id_col]],

+	other_raw = .data[[other_col]]

+	) %>%

+	mutate(

+	other_raw_chr = trimws(as.character(other_raw)),

+	other_submitted = suppressWarnings(as.numeric(other_raw_chr)),

+	other_submitted = ifelse(is.na(other_submitted), 0, other_submitted),

+	other_required = 2,

+	other_remaining = pmax(0, other_required - other_submitted),

+	other_complete = other_submitted >= other_required

+	)

+ } else {

+	other_df <- tracker %>%

+	transmute(

+	!!id_col := .data[[id_col]],

+	other_submitted = 0,

+	other_required = 2,

+	other_remaining = 2,

+	other_complete = FALSE

+	)

+ }

> > # ----------------------------

> # 15) Combine summary

> # ----------------------------

> summary_tbl <- fixed_summary %>%

+	left_join(other_df, by = id_col) %>%

+	mutate(

+	overall_complete = (n_complete == n_total_fixed) & other_complete

+	)

> > # ----------------------------

> # 16) Create R Markdown template

> # ----------------------------

> rmd_text <- c(

+	'---',

+	'title: "Waveform Portfolio Completion Report"',

+	'output:',

+	' pdf_document:',

+	'	latex_engine: pdflatex',

+	'params:',

+	' learner_name: ""',

+	' learner_email: ""',

+	' summary_table: !r data.frame()',

+	' missing_by_section: !r list()',

+	' text_only_by_section: !r list()',

+	' other_submitted: 0',

+	' other_required: 2',

+	' other_remaining: 2',

+	' other_complete: false',

+	'geometry: margin=1in',

+	'---',

+	'',

+	'```{r setup, include=FALSE}',

+	'knitr::opts_chunk$set(echo = FALSE, warning = FALSE, message = FALSE)',

+	'library(knitr)',

+	'',

+	'print_section_items <- function(section_name, item_list, empty_text) {',

+	' cat("\\n\\n## ", section_name, "\\n\\n", sep = "")',

+	' if (length(item_list) == 0) {',

+	'	cat(empty_text)',

+	' } else {',

+	'	cat(paste0("- ", unlist(item_list)), sep = "\\n")',

+	' }',

+	'}',

+	'```',

+	'',

+	'# Learner Summary',

+	'',

+	'**Name:** `r params$learner_name` ',

+	'**Email:** `r ifelse(is.null(params$learner_email) || is.na(params$learner_email) || params$learner_email == "NA" || params$learner_email == "", "", params$learner_email)`',

+	'',

+	'```{r}',

+	'kable(params$summary_table, booktabs = TRUE, align = c("l", "r"))',

+	'```',

+	'',

+	'# Missing Items',

+	'',

+	'```{r, results="asis"}',

+	'section_order <- c(',

+	' "Normal Waveforms",',

+	' "Maneuvers",',

+	' "Expiratory Asynchronies",',

+	' "Trigger Asynchronies",',

+	' "Flow Asynchronies",',

+	' "Cycle Asynchronies"',

+	')',

+	'',

+	'for (sec in section_order) {',

+	' items <- params$missing_by_section[[sec]]',

+	' if (is.null(items)) items <- list()',

+	' print_section_items(sec, items, "No items missing in this section.")',

+	'}',

+	'```',

+	'',

+	'# Text Entered But Image Still Needed',

+	'',

+	'```{r, results="asis"}',

+	'for (sec in section_order) {',

+	' items <- params$text_only_by_section[[sec]]',

+	' if (is.null(items)) items <- list()',

+	' print_section_items(sec, items, "No text-only items in this section.")',

+	'}',

+	'```',

+	'',

+	'# Other',

+	'',

+	'Submit **2 varied waveform examples**. Refer back to the portfolio instructions for acceptable submissions.',

+	'',

+	'```{r, results="asis"}',

+	'if (isTRUE(params$other_complete)) {',

+	' cat("Status: Complete (", params$other_submitted, " of ", params$other_required, " submitted).", sep = "")',

+	'} else {',

+	' cat("Status: Incomplete (", params$other_submitted, " of ", params$other_required, " submitted; ",',

+	'	params$other_remaining, " additional varied waveform",',

+	'	ifelse(params$other_remaining == 1, " is", "s are"), " needed).", sep = "")',

+	'}',

+	'```',

+	'',

+	'# Overall Status',

+	'',

+	'```{r, results="asis"}',

+	'any_missing <- any(lengths(params$missing_by_section) > 0)',

+	'any_text_only <- any(lengths(params$text_only_by_section) > 0)',

+	'',

+	'if (!any_missing && !any_text_only && isTRUE(params$other_complete)) {',

+	' cat("This learner\\\'s waveform portfolio is complete.")',

+	'} else {',

+	' cat("This learner still has outstanding portfolio requirements. Please complete the items listed above.")',

+	'}',

+	'```'

+ )

> > writeLines(rmd_text, template_file)

> > # ----------------------------

> # 17) Render one PDF per learner

> # ----------------------------

> section_order <- c(

+	"Normal Waveforms",

+	"Maneuvers",

+	"Expiratory Asynchronies",

+	"Trigger Asynchronies",

+	"Flow Asynchronies",

+	"Cycle Asynchronies"

+ )

> > for (i in seq_len(nrow(summary_tbl))) {

+

+	learner_name <- as.character(summary_tbl[[id_col]][i])

+	learner_email <- as.character(summary_tbl[[email_col]][i])

+

+	learner_missing <- grouped_details %>%

+	filter(.data[[id_col]] == learner_name, status == "missing")

+

+	learner_text_only <- grouped_details %>%

+	filter(.data[[id_col]] == learner_name, status == "text_only")

+

+	missing_by_section <- setNames(vector("list", length(section_order)), section_order)

+	text_only_by_section <- setNames(vector("list", length(section_order)), section_order)

+

+	for (sec in section_order) {

+	miss_items <- learner_missing %>%

+	filter(section == sec) %>%

+	pull(items)

+

+	txt_items <- learner_text_only %>%

+	filter(section == sec) %>%

+	pull(items)

+

+	missing_by_section[[sec]] <- unlist(miss_items, use.names = FALSE)

+	text_only_by_section[[sec]] <- unlist(txt_items, use.names = FALSE)

+	}

+

+	summary_table_out <- data.frame(

+	Metric = c(

+	"Completed fixed items",

+	"Text only; image still needed",

+	"Missing fixed items",

+	"Total fixed items",

+	"Percent complete (fixed items)",

+	"Other varied waveforms submitted",

+	"Other varied waveforms required"

+	),

+	Value = c(

+	summary_tbl$n_complete[i],

+	summary_tbl$n_text_only[i],

+	summary_tbl$n_missing[i],

+	summary_tbl$n_total_fixed[i],

+	paste0(summary_tbl$pct_complete_fixed[i], "%"),

+	summary_tbl$other_submitted[i],

+	summary_tbl$other_required[i]

+	),

+	stringsAsFactors = FALSE

+	)

+

+	output_file <- paste0(safe_filename(learner_name), "_report.pdf")

+

+	rmarkdown::render(

+	input = template_file,

+	output_file = output_file,

+	output_dir = output_dir,

+	params = list(

+	learner_name = learner_name,

+	learner_email = learner_email,

+	summary_table = summary_table_out,

+	missing_by_section = missing_by_section,

+	text_only_by_section = text_only_by_section,

+	other_submitted = summary_tbl$other_submitted[i],

+	other_required = summary_tbl$other_required[i],

+	other_remaining = summary_tbl$other_remaining[i],

+	other_complete = summary_tbl$other_complete[i]

+	),

+	envir = new.env(parent = globalenv()),

+	quiet = TRUE

+	)

+

+	message("Created: ", file.path(output_dir, output_file))

+ }

> > # ----------------------------

> # 10) Export summary CSV

> # ----------------------------

> summary_export <- summary_tbl %>%

+	mutate(

+	portfolio_status = ifelse(overall_complete, "Complete", "Incomplete")

+	) %>%

+	select(

+	all_of(c(id_col, email_col)),

+	n_complete,

+	n_text_only,

+	n_missing,

+	n_total_fixed,

+	pct_complete_fixed,

+	other_submitted,

+	other_required,

+	other_remaining,

+	portfolio_status

+	)

> > write_csv(summary_export, file.path(output_dir, "WPFall25_grouped_summary.csv"))

> > cat("\nDone.\n")

**Exam Autograding Code**

const FORM_ID = '';

const SS_ID = '';

const KEY_ID = "";

const RESULTS_ID = "";

const SUMMARY_ID = "";

const COMPARISON_ID = "";

const DEFAULT_MAX_POINTS = 10;

const DEFAULT_MIN_POINTS = -10;

const COL_LEARNER_ID = 1;

const COL_PRE_POST = 2;

const COL_QUESTIONS_START = 3;

const PRE_LABEL = "Pre";

const POST_LABEL = "Post";

function generateAnswerKey() {

const ss = SpreadsheetApp.getActiveSpreadsheet();

let sheet = ss.getSheetByName(KEY_ID);

if (sheet) {

SpreadsheetApp.getUi().alert("Answer Key already exists.");

return;

}

sheet = ss.insertSheet(KEY_ID);

const form = FormApp.openById(FORM_ID);

const builderHeaders = ['Question', 'Logic', 'Answer', 'Unless Selected', 'Points', 'Max Points', 'Min Points'];

sheet.getRange(1, 1, 1, 7).setValues([builderHeaders]);

const qTitles = form.getItems()

.filter(item => item.getType() === FormApp.ItemType.CHECKBOX)

.map(item => item.getTitle());

const qValidation = SpreadsheetApp.newDataValidation().requireValueInList(qTitles).build();

const logicValidation = SpreadsheetApp.newDataValidation().requireValueInList(['BOTH', 'EITHER', 'EACH']).build();

sheet.getRange(2, 1, 500, 1).setDataValidation(qValidation);

sheet.getRange(2, 2, 500, 1).setDataValidation(logicValidation);

}

function scrapeAnswerKey() {

const ss = SpreadsheetApp.getActiveSpreadsheet();

const sheet = ss.getSheetByName(KEY_ID);

const data = sheet.getRange(2, 1, sheet.getLastRow(), 7).getValues();

let answerKeyMap = {};

let pointLimitMap = {};

const priorityMap = { "BOTH": 1, "EITHER": 2, "EACH": 2 };

for (let i = 0; i < data.length; i++) {

let [q, logic, choices, unless, points, maxPts, minPts] = data[i];

if (!q || !logic) continue;

if (!answerKeyMap[q]) answerKeyMap[q] = [];

answerKeyMap[q].push({

logic: String(logic).toUpperCase(),

options: String(choices).split(",").map(s => s.trim()),

unless: String(unless).split(",").map(s => s.trim()).filter(s => s !== ""),

points: Number(points) || 0,

priority: priorityMap[String(logic).toUpperCase()] || 3

});

if (maxPts !== "" && !isNaN(maxPts)) {

pointLimitMap[q] = {

max: Number(maxPts),

min: (minPts !== "" && !isNaN(minPts)) ? Number(minPts) : DEFAULT_MIN_POINTS

};

}

}

for (let q in answerKeyMap) {

answerKeyMap[q].sort((a, b) => a.priority - b.priority || b.options.length - a.options.length);

}

return { answerKeyMap, pointLimitMap };

}

function gradeExam(studentAnswers, answerKeyMap, pointLimitMap) {

let grandTotal = 0;

let questionBreakdown = {};

for (let q in answerKeyMap) {

let qSubtotal = 0;

let selected = studentAnswers[q] || [];

let remainingTokens = new Set(selected);

let rules = answerKeyMap[q];

rules.forEach(rule => {

if (rule.unless.some(u => selected.includes(u))) return;

if (rule.options.includes("[BLANK]")) {

if (selected.length === 0) qSubtotal += rule.points;

return;

}

let matches = rule.options.filter(opt => remainingTokens.has(opt));

if (rule.logic === "BOTH" && matches.length === rule.options.length && rule.options.length > 0) {

qSubtotal += rule.points;

rule.options.forEach(opt => remainingTokens.delete(opt));

} else if (rule.logic === "EITHER" && matches.length > 0) {

qSubtotal += rule.points;

matches.forEach(opt => remainingTokens.delete(opt));

} else if (rule.logic === "EACH") {

qSubtotal += (matches.length * rule.points);

matches.forEach(opt => remainingTokens.delete(opt));

}

});

let limits = pointLimitMap[q];

let fMax = (limits && limits.max !== undefined) ? limits.max : DEFAULT_MAX_POINTS;

let fMin = (limits && limits.min !== undefined) ? limits.min : DEFAULT_MIN_POINTS;

if (qSubtotal > fMax) qSubtotal = fMax;

if (qSubtotal < fMin) qSubtotal = fMin;

questionBreakdown[q] = qSubtotal;

grandTotal += qSubtotal;

}

return { total: grandTotal, breakdown: questionBreakdown };

}

function parseResponseRow(rowData, headers, checkboxTitles, itemAnalysis) {

let studentAnswers = {};

let nonCheckboxAnswers = {};

for (let j = COL_QUESTIONS_START; j < headers.length; j++) {

let qTitle = headers[j];

let val = rowData[j];

if (checkboxTitles.includes(qTitle)) {

let rawChoices = String(val).split(",").map(s => s.trim()).filter(Boolean);

if (itemAnalysis && itemAnalysis[qTitle]) {

rawChoices.forEach(choice => {

itemAnalysis[qTitle].frequencies[choice] = (itemAnalysis[qTitle].frequencies[choice] || 0) + 1;

});

}

studentAnswers[qTitle] = rawChoices.map(r => String(r).split(/[.\s]/)[0].trim());

} else {

nonCheckboxAnswers[qTitle] = val;

}

}

return { studentAnswers, nonCheckboxAnswers };

}

function buildComparisonSheet(preGrades, postGrades, questionHeaders, checkboxTitles) {

const ss = SpreadsheetApp.getActiveSpreadsheet();

let sheet = ss.getSheetByName(COMPARISON_ID) || ss.insertSheet(COMPARISON_ID);

sheet.clear();

const allLearners = [...new Set([...Object.keys(preGrades), ...Object.keys(postGrades)])].sort();

const qCols = questionHeaders.filter(q => checkboxTitles.includes(q));

let headers = ['Learner ID', 'Pre Total', 'Post Total', 'Total Diff'];

qCols.forEach(q => headers.push(`${q} — Pre`, `${q} — Post`, `${q} — Diff`));

sheet.appendRow(headers);

let rows = [];

allLearners.forEach(learnerId => {

const pre = preGrades[learnerId] || null;

const post = postGrades[learnerId] || null;

const preTotal = pre ? pre.total : '';

const postTotal = post ? post.total : '';

const totalDiff = (pre && post) ? (post.total - pre.total) : '';

let row = [learnerId, preTotal, postTotal, totalDiff];

qCols.forEach(q => {

const preQ = (pre && pre.breakdown[q] !== undefined) ? pre.breakdown[q] : '';

const postQ = (post && post.breakdown[q] !== undefined) ? post.breakdown[q] : '';

const diff = (preQ !== '' && postQ !== '') ? (postQ - preQ) : '';

row.push(preQ, postQ, diff);

});

rows.push(row);

});

if (rows.length > 0) {

sheet.getRange(2, 1, rows.length, headers.length).setValues(rows);

}

const statsStartRow = rows.length + 3;

const preScores = allLearners.map(id => preGrades[id] ? preGrades[id].total : null).filter(v => v !== null);

const postScores = allLearners.map(id => postGrades[id] ? postGrades[id].total : null).filter(v => v !== null);

const pairedDiffs = allLearners

.filter(id => preGrades[id] && postGrades[id])

.map(id => postGrades[id].total - preGrades[id].total);

const avg = arr => arr.length > 0 ? (arr.reduce((a, b) => a + b, 0) / arr.length).toFixed(2) : '';

sheet.getRange(statsStartRow, 1).setValue('Overall Stats');

sheet.getRange(statsStartRow + 1, 1, 5, 3).setValues([

['', PRE_LABEL, POST_LABEL],

['Count', preScores.length, postScores.length],

['Average Total Score', avg(preScores), avg(postScores)],

['Paired learners', pairedDiffs.length, pairedDiffs.length],

['Average Score Change (paired)', avg(pairedDiffs), ''],

]);

}

function manualRegrade() {

const ss = SpreadsheetApp.getActiveSpreadsheet();

const ui = SpreadsheetApp.getUi();

const respSheet = ss.getSheetByName("Form Responses");

if (!respSheet) return;

const existing = ss.getSheetByName(RESULTS_ID);

if (existing) {

const confirm = ui.alert(

'Overwrite existing results?',

'The Grades, Summary Stats, and Pre/Post Comparison sheets will be cleared and rebuilt. This cannot be undone.',

ui.ButtonSet.YES_NO

);

if (confirm !== ui.Button.YES) return;

}

const lock = LockService.getScriptLock();

try {

lock.waitLock(30000);

} catch (e) {

ui.alert('Another grading run is in progress. Please wait and try again.');

return;

}

try {

const form = FormApp.openById(FORM_ID);

const checkboxTitles = form.getItems(FormApp.ItemType.CHECKBOX).map(item => item.getTitle());

const { answerKeyMap, pointLimitMap } = scrapeAnswerKey();

const data = respSheet.getDataRange().getValues();

const headers = data[0];

const questionHeaders = headers.slice(COL_QUESTIONS_START);

let resultsSheet = ss.getSheetByName(RESULTS_ID) || ss.insertSheet(RESULTS_ID);

resultsSheet.clear();

const newHeaders = ['Timestamp', 'Learner ID', 'Pre/Post', ...questionHeaders, 'Final Score', 'Duplicate'];

resultsSheet.appendRow(newHeaders);

let results	= [];

let preScoreList = [], postScoreList = [];

let preAnalysis = {}, postAnalysis = {};

let preGrades	= {}, postGrades	= {};

let preSeenIds	= {}, postSeenIds	= {};

let duplicateFlags = {};

questionHeaders.forEach(q => {

if (checkboxTitles.includes(q)) {

let qMax = (pointLimitMap[q] && pointLimitMap[q].max !== undefined) ? pointLimitMap[q].max : DEFAULT_MAX_POINTS;

preAnalysis[q] = { totalPoints: 0, fullCreditCount: 0, frequencies: {}, customMax: qMax };

postAnalysis[q] = { totalPoints: 0, fullCreditCount: 0, frequencies: {}, customMax: qMax };

}

});

for (let i = 1; i < data.length; i++) {

const row	= data[i];

const learnerId = String(row[COL_LEARNER_ID]).trim();

const prePost	= String(row[COL_PRE_POST]).trim();

const isPost	= prePost === POST_LABEL;

const analysis = isPost ? postAnalysis : preAnalysis;

const { studentAnswers, nonCheckboxAnswers } = parseResponseRow(row, headers, checkboxTitles, analysis);

const gradingResults = gradeExam(studentAnswers, answerKeyMap, pointLimitMap);

let isDuplicate = false;

if (isPost) {

postSeenIds[learnerId] = (postSeenIds[learnerId] || 0) + 1;

if (postSeenIds[learnerId] > 1) {

isDuplicate = true;

if (!duplicateFlags[learnerId]) duplicateFlags[learnerId] = [];

if (!duplicateFlags[learnerId].includes(POST_LABEL)) duplicateFlags[learnerId].push(POST_LABEL);

}

postGrades[learnerId] = gradingResults;

postScoreList.push(gradingResults.total);

} else {

preSeenIds[learnerId] = (preSeenIds[learnerId] || 0) + 1;

if (preSeenIds[learnerId] > 1) {

isDuplicate = true;

if (!duplicateFlags[learnerId]) duplicateFlags[learnerId] = [];

if (!duplicateFlags[learnerId].includes(PRE_LABEL)) duplicateFlags[learnerId].push(PRE_LABEL);

}

preGrades[learnerId] = gradingResults;

preScoreList.push(gradingResults.total);

}

let gradeRow = [row[0], learnerId, prePost];

questionHeaders.forEach(qTitle => {

if (checkboxTitles.includes(qTitle)) {

let pts = gradingResults.breakdown[qTitle] || 0;

gradeRow.push(pts);

analysis[qTitle].totalPoints += pts;

if (pts >= analysis[qTitle].customMax) analysis[qTitle].fullCreditCount += 1;

} else {

gradeRow.push(nonCheckboxAnswers[qTitle] || "");

}

});

gradeRow.push(gradingResults.total);

gradeRow.push(isDuplicate ? 'DUPLICATE' : '');

results.push(gradeRow);

}

if (results.length > 0) {

resultsSheet.getRange(2, 1, results.length, newHeaders.length).setValues(results);

}

let summarySheet = ss.getSheetByName(SUMMARY_ID) || ss.insertSheet(SUMMARY_ID);

summarySheet.clear();

function appendCohortStats(scores, itemAnalysis, label) {

const count	= scores.length;

const sum	= scores.reduce((a, b) => a + b, 0);

const avgFinal = count > 0 ? (sum / count).toFixed(2) : 0;

summarySheet.appendRow([label]);

summarySheet.appendRow(['Average Final Score', avgFinal]);

summarySheet.appendRow(['Total Students', count]);

summarySheet.appendRow([' ']);

for (let q in itemAnalysis) {

let stats	= itemAnalysis[q];

let avgScore = count > 0 ? (stats.totalPoints / count).toFixed(2) : 0;

let fullCreditRate = count > 0 ? ((stats.fullCreditCount / count) * 100).toFixed(1) + "%" : "0%";

summarySheet.appendRow([q, 'Average Score:', avgScore, 'Full Credit Rate:', fullCreditRate]);

summarySheet.appendRow(['', 'Option Selected', 'Count', '% of Class']);

let sortedChoices = Object.entries(stats.frequencies).sort((a, b) => b[1] - a[1]);

if (sortedChoices.length === 0) {

summarySheet.appendRow(['', 'No answers recorded', '0', '0%']);

} else {

sortedChoices.forEach(([choice, cCount]) => {

summarySheet.appendRow(['', choice, cCount, ((cCount / count) * 100).toFixed(1) + "%"]);

});

}

summarySheet.appendRow([' ']);

}

summarySheet.appendRow([' ']);

}

appendCohortStats(preScoreList, preAnalysis, '── Pre-Test ──');

appendCohortStats(postScoreList, postAnalysis, '── Post-Test ──');

buildComparisonSheet(preGrades, postGrades, questionHeaders, checkboxTitles);

ss.getSheetByName(COMPARISON_ID).activate();

const dupIds = Object.keys(duplicateFlags);

if (dupIds.length > 0) {

const dupLines = dupIds.map(id => `${id}: duplicate ${duplicateFlags[id].join(' & ')} submission`);

ui.alert(

'Grading complete — duplicates detected',

`The following learners had multiple submissions for the same side. The last submission was used for grading and comparison.\n\n${dupLines.join('\n')}\n\nRows are marked DUPLICATE in the Grades sheet.`,

ui.ButtonSet.OK

);

} else {

ui.alert('Grading complete');

}

} finally {

lock.releaseLock();

}

}

function onOpen() {

SpreadsheetApp.getUi().createMenu('Dashboard')

.addItem('Generate Answer Key', 'generateAnswerKey')

.addItem('Regrade', 'manualRegrade')

.addToUi();

}
